# Deep vein thrombosis in critically ill patients with COVID-19: Incidence, Wells score diagnosis validation, and hospital prognosis

**DOI:** 10.1101/2025.04.30.25326684

**Authors:** Clévio Cezar da Fonseca, Hugo Perazzo Pedroso Barbosa, Sandra Wagner Cardoso, Isabel Cristina Ferreira Tavares, Maria Pia Diniz Ribeiro, Rodrigo de Carvalho Moreira, Lara Esteves Coelho, Emília Moreira Jalil, André Miguel Japiassú, Elias Pimentel Gouvêa, Estevão Portela Nunes, Hugo Boechat Andrade, Luciano Barros Gouvêa, Marcel Treptow Ferreira, Pedro Mendes Azambuja Rodrigues, Ronaldo Ismerio Moreira, Kim Mattos Geraldo, Lucilene Araújo de Freitas, Vinicius Velleda Pacheco, Beatriz Gilda Jegerhorn Grinsztein, Pedro Emmanuel Alvarenga Americano do Brasil

## Abstract

**BACKGROUND:** The risk of major venous thromboembolism (VTE) among patients with COVID-19 is high but varies with disease severity.

**OBJECTIVE:** Estimate the incidence of lower extremity deep venous thrombosis (DVT) in critically ill hospitalized patients with COVID-19, validate the Wells score for DVT diagnosis, and determine patient’s prognosis.

**DESIGN AND SETTING:** This was an observational follow-up study in the context of diagnosis and prognosis of DVT. All patients hospitalized in the intensive care unit (ICU) of the Evandro Chagas National Institute of Infectious Diseases.

**METHODS:** Participants with COVID-19 pneumonia were included. Lower-limb Doppler to assess DVT was performed at admission and follow-up. Prognosis outcomes were death, length of stay, need for mechanical ventilation, vasopressor use, and hemodialysis. Wells’ score sensitivity and specificity were estimated at admission. Survival curves were estimated for patients with DVT and adjusted for the SAPS3 score.

**RESULTS:** Between June 2020 and January 2021, 186 patients were included. The DVT incidence was 0.097. A Wells score of two or higher had a sensitivity and specificity of 1.00 and 0.94 respectively. Mortality and mechanical ventilation support were higher in participants with DVT. For these outcomes, after SAPS 3 adjustment, participants with DVT had twice the hazard of those without DVT. A web calculator (https://pedrobrasil.shinyapps.io/INDWELL/) is available for predictions.

**CONCLUSIONS:** One can use the Wells score to accurately diagnose DVT in critically ill patients with COVID-19. DVT increases severity of COVID-19, which highlights the importance of its early diagnosis, treatment, and prophylaxis at the ICU setting.

## BACKGROUND

By October 2022, 617,787,221 Coronavirus disease 2019 (COVID-19) cases and 6,545,929 deaths due to COVID-19 were confirmed worldwide. Brazil has recorded 34,678,510 cases and 686,254 deaths in the same period.^1^ From 10.96% to 22.90% COVID-19 cases may progress requiring hospitalization in Intensive Care Units (ICU).^2,3^ The overall COVID-19 mortality in critically ill patients is variable ranging from 5.6%^3^ to 49%,^4^ with the length of stay ranging from few days to several weeks.^5^ Additionally, the estimated risk of mechanical ventilation was estimated to be 7.1%.^3^

Coagulation disturbances are frequent clinical manifestations of COVID-19 with critical illness, such as thrombocytopenia and elevated D-dimer due to abnormalities in the clotting cascade and are related to deep vein thrombosis (DVT) and mortality.^4^ Studies with Italian and Spanish populations showed DVT incidences among critically ill COVID-19 patients ranging from 7.7% to 46%.^2,6–8^ Studies at Brazil demonstrated a DVT incidence ranging from 1.6% to 36.8 % among critically ill COVID-19 patients.^9–12^ The risk of major venous thromboembolism (VTE) among patients with COVID-19 is high, but varies with disease severity.^13^ The mortality in critically ill patients with COVID-19 with DVT ranges from 11.4 to 70.0%.^10,12,14,15^ Severe acute respiratory syndrome coronavirus 2 (SARS-CoV-2) may directly cause endothelial injury, which may also impact DVT incidence. The mechanism by which COVID-19 increases the risk of acute pulmonary thromboembolism (PTE) is not fully understood.^16^

The VTE incidence was estimated to be one case per 1,000 inhabitants per year, and this nosological categorization was responsible for the death of approximately 50,000 North Americans each year.^17^ VTE is a manifestation of DVT, which manifests commonly as PTE, with or without detectable DVT.^18,19^ Almost two-thirds of VTE cases are associated with a single DVT finding, and the majority of DVT cases occur in the lower limbs. VTE is the third most life-threatening cause of acute cardiovascular syndromes worldwide. Approximately 34% of VTE-affected patients die suddenly.^19^

In 1997, Wells^20^ developed a clinical score to conduct diagnostic investigations and classify patients according to their DVT risk. This score is currently used and recommended by guidelines^19,21^ to estimate DVT diagnosis probability before performing ultrasonography (US).

Twenty percent of distal DVT will progress to proximal thrombosis, increasing the risk of death. Therefore, identifying the DVT location and classifying its type is crucial for determining the treatment strategies.^22^ The Brazilian Society Angiology and Vascular Surgery (SBACV) Guidelines Project (2015) outlines detailed diagnostic and treatment recommendations for different DVT locations and classifications.^22^

Efforts have been made to establish whether intensified thromboprophylaxis regimens are required for critically ill patients with COVID-19. The best thromboprophylaxis should balance the risks of thrombosis and bleeding. However, studies on VTE with COVID-19 in Brazil are scarce. Most studies on this topic have been conducted on European and North American populations. The American Society of Hematology guidelines on thromboprophylaxis in patients with COVID-19 stated the importance of an individualized decision for each patient based on an assessment of thrombosis and bleeding risk.^23^ Therefore, timely assessment of DVT and implementation of preventive strategies are necessary for a favorable prognosis of critically ill patients with COVID-19.^24^

## OBJECTIVES

This study aims are to estimate the DVT incidence in patients with COVID-19 hospitalized in a critical care unit. This study also aims to validate the Wells score for DVT diagnosis among critically ill patients with COVID-19 and to estimate the prognosis of patients with COVID-19 and DVT.

## METHODS

DVT diagnosis and prognosis in patients with COVID-19 is a RECOVER-SUS [NCT04807699] sub-study. RECOVER-SUS is a prospective observational multicenter follow-up study of hospitalized patients with COVID-19 performed in seven public tertiary hospitals across five cities in Brazil.^25^ The DVT sub-study was conducted at Instituto Nacional de Infectologia Evandro Chagas (INI/FIOCRUZ) only. INI/FIOCRUZ is part of the public health system. INI/FICORUZ is specialized in infectious diseases health care. At the time of the COVID-19 pandemic it had between 140 and 200 beds, including intermediate and ICU beds dedicated to CVOID-19 care. INI/FIOCRUZ admitted patients transferred from other health units, emergency rooms, from primary and intermediate complexity and from neighboring cities.

Patients eligible for participation in the RECOVER-SUS study included critically hospitalized patients, aged 18 years or older, with evidence of SARS-CoV-2 infection as per World Health Organization (WHO) criteria^26^ within 14 days of the onset of symptoms initiation, confirmed through PCR testing of nasopharyngeal, oropharyngeal, or tracheal aspirate samples. These individuals also needed to exhibit chest imaging abnormalities at the time of screening. Participants without suspected, probable, or confirmed SARS-CoV-2 infection according to the WHO COVID-19 guidelines were excluded.

This sub-study included a cross-sectional diagnostic study at the time of ICU admission and an observational follow-up prognostic study developed in one of the ICUs (ICU-H unit). During the study period, the ICU was exclusively for COVID-19 patients. All patients hospitalized in ICU-H during the inclusion period were sequentially screened for the sub-study. In addition to the RECOVER-SUS criteria, the sub-study inclusion criteria included indication for ICU admission due to COVID-19-related respiratory complications, such as oxygen saturation ≤94% or the requirement for supplemental oxygen (including non-invasive positive pressure ventilation or high flow supplemental oxygen), invasive mechanical ventilation (IMV), or extracorporeal membrane oxygenation (ECMO), and the performance of deep vein ultrasonography (US) in the first 48 h of admission. The screening period for the DVT sub-study was from June 2020 to January 2021. The DVT sub-study has a retrospective part, checking and extracting data from June to October 2020, and a prospective part going along with the original RECOVER-SUS effort up to the end of the data collection period. Vaccination in Brazil began on January 17, 2021; therefore, all participants were not vaccinated against COVID-19.^27^

Socio-demographic characteristics, comorbidities, COVID-19 symptoms, vital signs, and anthropometric measurements (weight and height) were recorded at hospital admission (baseline). Clinical data, blood samples, and ultrasound images for DVT verification (identifying the incidence and sites of location) were collected by trained investigators at baseline and on days 3, 7, 14, 21, and 28 of hospitalization. The study data were collected and managed using REDCap electronic data capture tool hosted at INI/FIOCRUZ. All participants were monitored continuously from the time of hospital admission until they were either transferred to another institution, discharged, deceased or until the sub-study was interrupted.

The severity scores and Wells’ score were obtained upon ICU admission. The Wells’ score questions pertained to the incidence of active cancer treatment or palliation within six months); history of being bedridden recently either for more than three days or the occurrence of major surgery within 12 weeks; calf swelling in one leg measuring more than 3 cm in comparison to the other leg, with measurements taken 10 cm below the tibial tuberosity; the presence of collateral (non-varicose) superficial veins; swelling in the entire leg; localized tenderness along the deep venous system; pitting edema, confined to symptomatic leg; paralysis, paresis, or recent plaster immobilization of the lower extremity; previously documented DVT alternative diagnosis, as likely or more likely. The Wells score ranges from 2 to 9, the higher the score more likely the DVT diagnosis.^28,29^ The Charlson Comorbidity Index (CCI)^30^, the Sequential Organ Failure Assessment (SOFA)^31^, the Simplified Acute Physiology Score version 3 (SAPS3)^32–35^, and the sepsis-induced coagulopathy (SIC)^36^ are usually used to predict mortality either in general, or due to comorbidities, organ failure or sepsis. These scores allow prognosis estimation adjustments

Point-of-care US DVT was assessed by critical care physician trained in point-of-care ultrasound (POCUS). The protocol to perform the test was to begin at the common femoral vein and move distally, repeating the procedure on the greater saphenous vein, always compressing at 1–2 cm intervals until the vessel was no longer visible. The protocol included visualization of the entire proximal superficial femoral vein using a standard linear probe. Finally, the popliteal vein was compressed.^37^ The equipment available to perform POCUS DVT was Viamo™ c100, a portable US system of a cart-based machine.^38^ For the validation of the Wells score, the US was used as the reference test. For prognostic purposes, the following outcomes were considered, including death, length of stay, need for mechanical ventilation, vasopressor use, and hemodialysis.

The DVT sub-study sample size was determined by the hospitalization capacity of the intensive care unit during the predetermined period of inclusion, which at the time was believed to be between 180 and 250. Baseline clinical characteristics were tabulated to show either their frequencies or central tendencies according to their format. Regarding the diagnostic aspect, we estimated the sensitivity and specificity for the Wells’ (and other) scores along with their 95% confidence intervals with binomial distribution with the Wilson method, with the actual score being considered as a positive test result. For this analysis, DVT at US at any moment of hospitalization was the reference test. Regarding the prognostic aspect, the DVT US diagnosis was adjusted with the SAPS 3 score using either a Cox proportional hazards model or a linear model, depending on the prognosis being considered. The time-dependent survival approach was used to consider participants who developed DVT either at baseline or during the follow-up period. The Wald test at 5% was used for significance.

## RESULTS

During the study period, 229 participants were screened. Of these, forty-three were excluded because they were not subjected to US within the first 48 h. A total of 186 patients were included in this analysis. DVT occurred in eighteen participants, consequently, the incidence and its respective 95% confidence interval were determined to be 0.097 [0.062–0.148]. Of these, ten participants were diagnosed with DVT within the first 24 h of admission, one participant was diagnosed after three days, while six participants were diagnosed after seven days, and one patient was diagnosed after 21 days of admission.

Age, sex, and ethnicity were similar between the participants within the DVT groups. Additionally, there were no evident differences in the DVT groups concerning the history of exposure, duration of symptoms, or physiological variables. (Table 1). Pre-existing comorbidities that were more frequent among the participants included hypertension, diabetes, and obesity. All these three comorbidities occurred more frequently among participants without DVT. (Table 2)

**Table 1:**
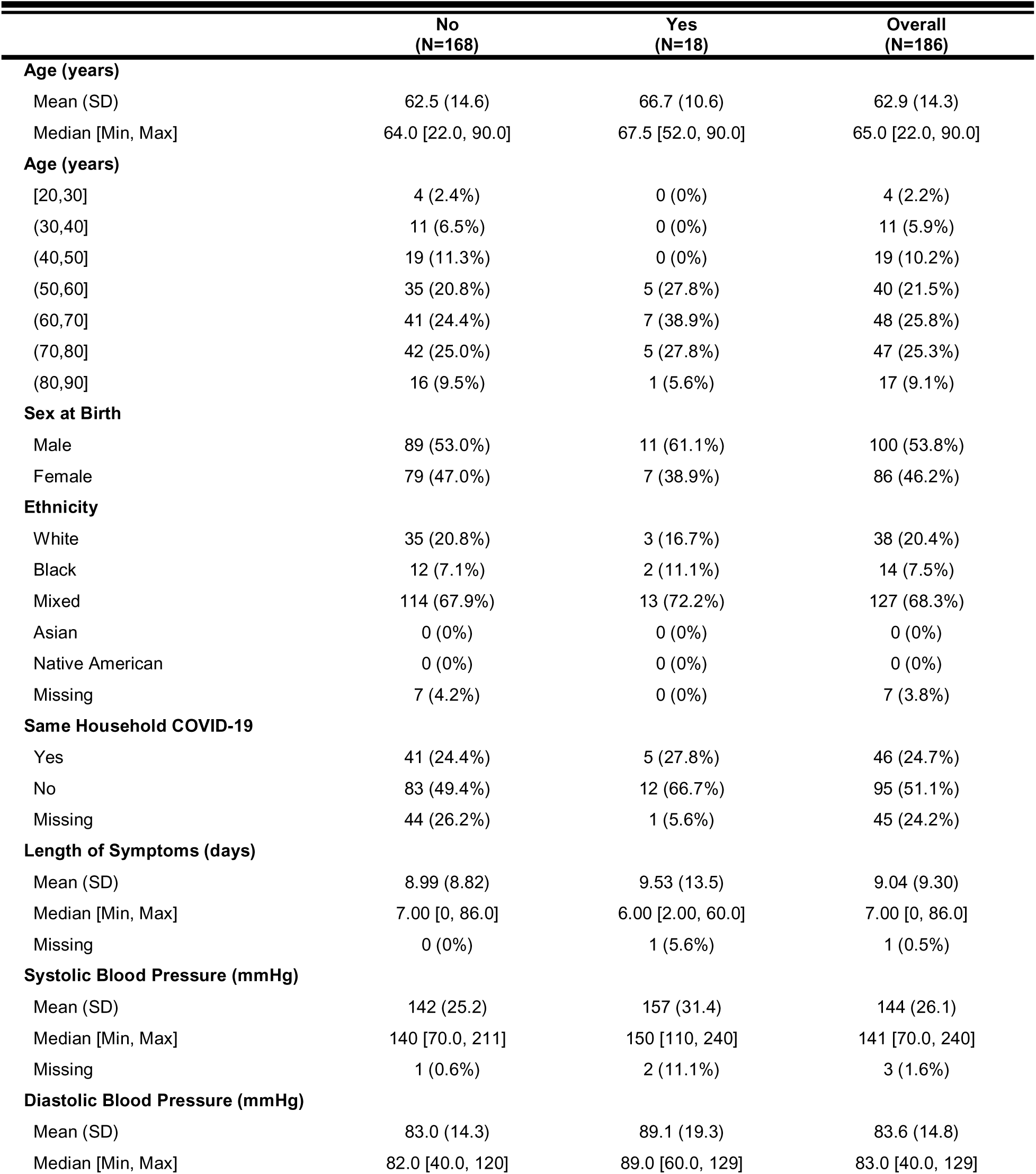
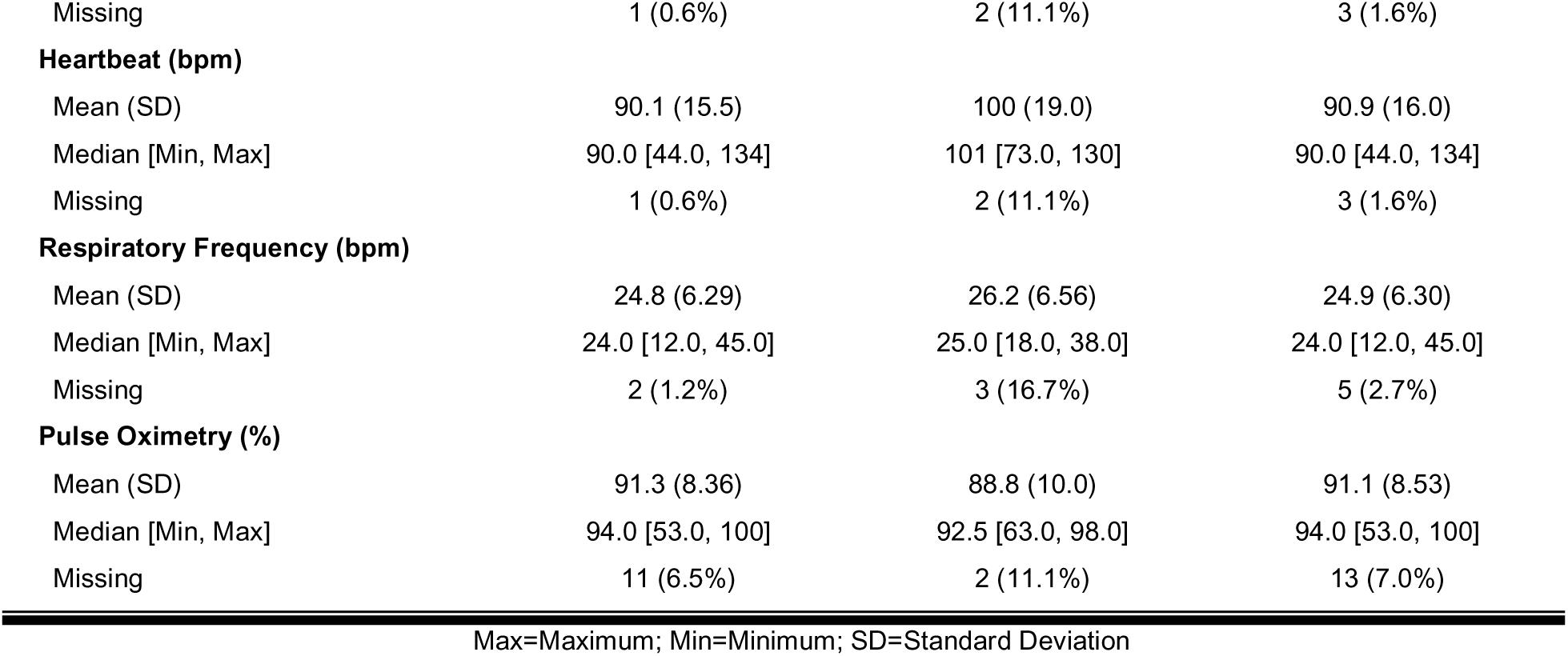
Clinical and demographic baseline data by deep venous thrombosis diagnosis.

**Table 2:**
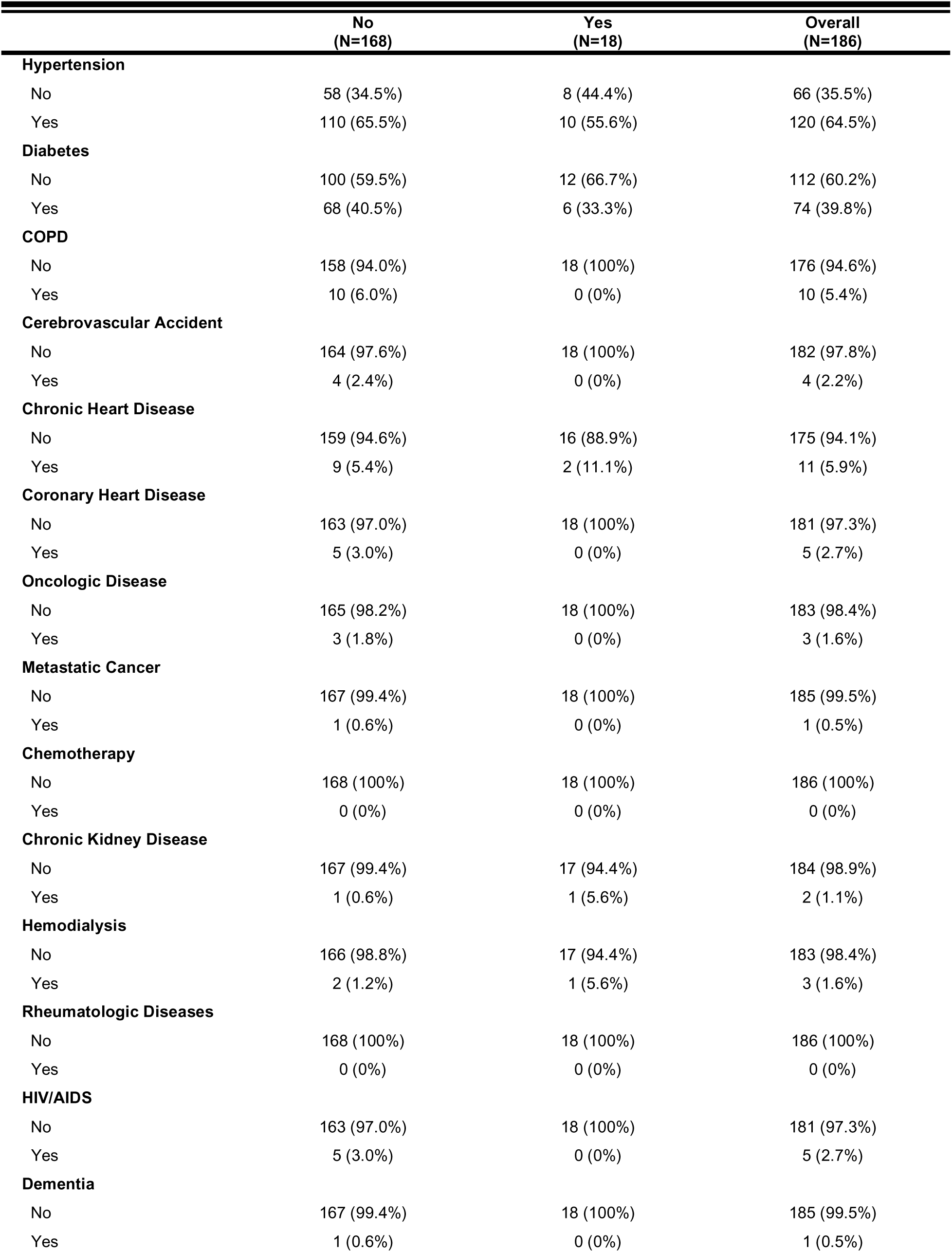
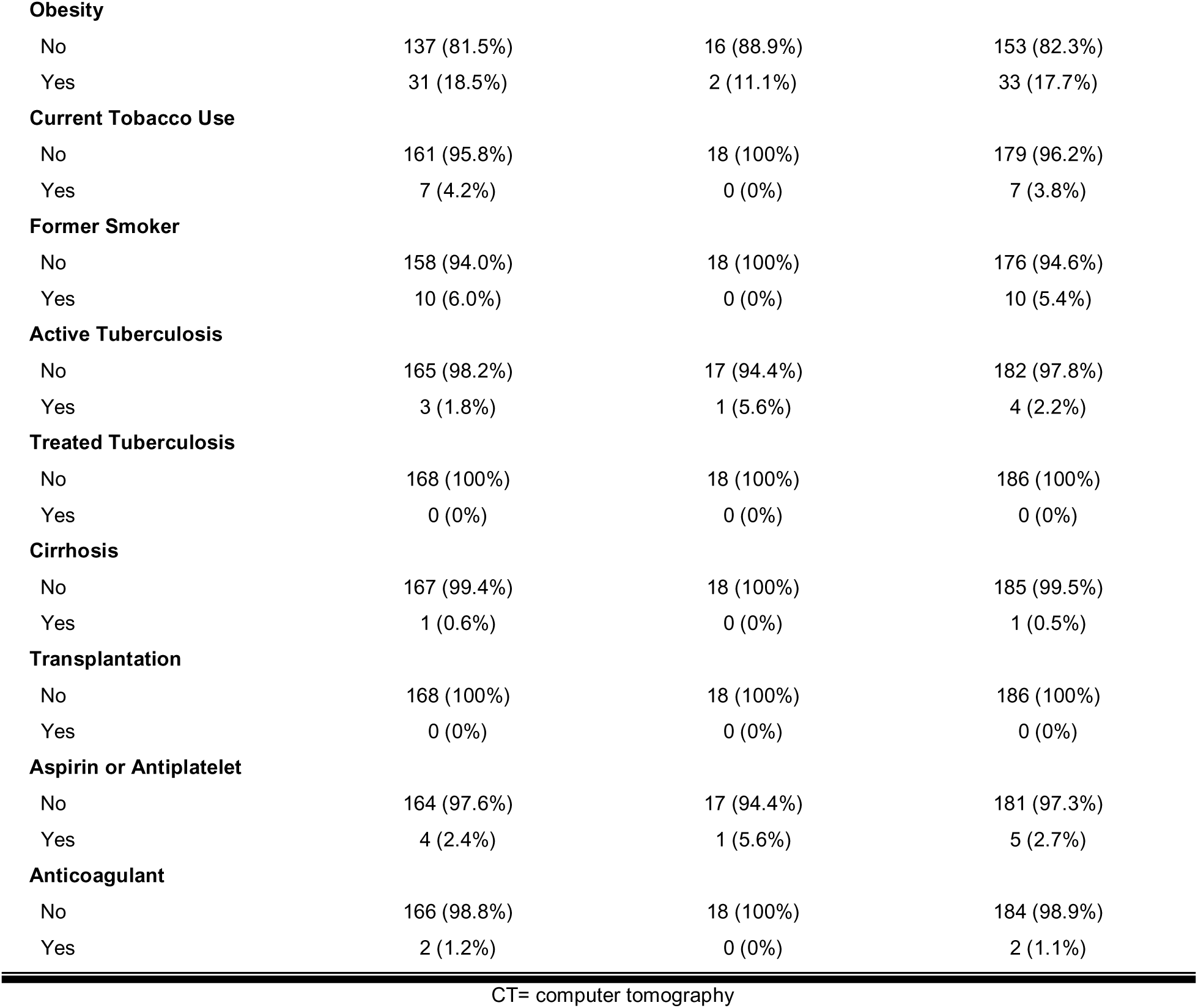
Comorbidities baseline data by deep venous thrombosis diagnosis.

|  | No<br>(N=168) | Yes<br>(N=18) | Overall<br>(N=186) |
| --- | --- | --- | --- |
| **Hypertension** |  |  |  |
| No | 58 (34.5%) | 8 (44.4%) | 66 (35.5%) |
| Yes | 110 (65.5%) | 10 (55.6%) | 120 (64.5%) |
| **Diabetes** |  |  |  |
| No | 100 (59.5%) | 12 (66.7%) | 112 (60.2%) |
| Yes | 68 (40.5%) | 6 (33.3%) | 74 (39.8%) |
| **COPD** |  |  |  |
| No | 158 (94.0%) | 18 (100%) | 176 (94.6%) |
| Yes | 10 (6.0%) | 0 (0%) | 10 (5.4%) |
| **Cerebrovascular Accident** |  |  |  |
| No | 164 (97.6%) | 18 (100%) | 182 (97.8%) |
| Yes | 4 (2.4%) | 0 (0%) | 4 (2.2%) |
| **Chronic Heart Disease** |  |  |  |
| No | 159 (94.6%) | 16 (88.9%) | 175 (94.1%) |
| Yes | 9 (5.4%) | 2 (11.1%) | 11 (5.9%) |
| **Coronary Heart Disease** |  |  |  |
| No | 163 (97.0%) | 18 (100%) | 181 (97.3%) |
| Yes | 5 (3.0%) | 0 (0%) | 5 (2.7%) |
| **Oncologic Disease** |  |  |  |
| No | 165 (98.2%) | 18 (100%) | 183 (98.4%) |
| Yes | 3 (1.8%) | 0 (0%) | 3 (1.6%) |
| **Metastatic Cancer** |  |  |  |
| No | 167 (99.4%) | 18 (100%) | 185 (99.5%) |
| Yes | 1 (0.6%) | 0 (0%) | 1 (0.5%) |
| **Chemotherapy** |  |  |  |
| No | 168 (100%) | 18 (100%) | 186 (100%) |
| Yes | 0 (0%) | 0 (0%) | 0 (0%) |
| **Chronic Kidney Disease** |  |  |  |
| No | 167 (99.4%) | 17 (94.4%) | 184 (98.9%) |
| Yes | 1 (0.6%) | 1 (5.6%) | 2 (1.1%) |
| **Hemodialysis** |  |  |  |
| No | 166 (98.8%) | 17 (94.4%) | 183 (98.4%) |
| Yes | 2 (1.2%) | 1 (5.6%) | 3 (1.6%) |
| **Rheumatologic Diseases** |  |  |  |
| No | 168 (100%) | 18 (100%) | 186 (100%) |
| Yes | 0 (0%) | 0 (0%) | 0 (0%) |
| **HIV/AIDS** |  |  |  |
| No | 163 (97.0%) | 18 (100%) | 181 (97.3%) |
| Yes | 5 (3.0%) | 0 (0%) | 5 (2.7%) |
| **Dementia** |  |  |  |
| No | 167 (99.4%) | 18 (100%) | 185 (99.5%) |
| Yes | 1 (0.6%) | 0 (0%) | 1 (0.5%) |
| **Obesity** |  |  |  |
| No | 137 (81.5%) | 16 (88.9%) | 153 (82.3%) |
| Yes | 31 (18.5%) | 2 (11.1%) | 33 (17.7%) |
| **Current Tobacco Use** |  |  |  |
| No | 161 (95.8%) | 18 (100%) | 179 (96.2%) |
| Yes | 7 (4.2%) | 0 (0%) | 7 (3.8%) |
| **Former Smoker** |  |  |  |
| No | 158 (94.0%) | 18 (100%) | 176 (94.6%) |
| Yes | 10 (6.0%) | 0 (0%) | 10 (5.4%) |
| **Active Tuberculosis** |  |  |  |
| No | 165 (98.2%) | 17 (94.4%) | 182 (97.8%) |
| Yes | 3 (1.8%) | 1 (5.6%) | 4 (2.2%) |
| **Treated Tuberculosis** |  |  |  |
| No | 168 (100%) | 18 (100%) | 186 (100%) |
| Yes | 0 (0%) | 0 (0%) | 0 (0%) |
| **Cirrhosis** |  |  |  |
| No | 167 (99.4%) | 18 (100%) | 185 (99.5%) |
| Yes | 1 (0.6%) | 0 (0%) | 1 (0.5%) |
| **Transplantation** |  |  |  |
| No | 168 (100%) | 18 (100%) | 186 (100%) |
| Yes | 0 (0%) | 0 (0%) | 0 (0%) |
| **Aspirin or Antiplatelet** |  |  |  |
| No | 164 (97.6%) | 17 (94.4%) | 181 (97.3%) |
| Yes | 4 (2.4%) | 1 (5.6%) | 5 (2.7%) |
| **Anticoagulant** |  |  |  |
| No | 166 (98.8%) | 18 (100%) | 184 (98.9%) |
| Yes | 2 (1.2%) | 0 (0%) | 2 (1.1%) |

CT= computer tomography

None of the patients had a normal chest radiograph at admission and only one had a normal CT scan. The most frequent tomographic images were consolidation, ground-glass, and pleural effusion, and there were no evident differences among the DVT groups in these findings. (Table 3) D-dimer, procalcitonin, interleukin six, and C-reactive protein levels were generally elevated in all participants. The D-dimer level was the only biomarker that was significantly higher in the DVT group (Table 4).

**Table 3:** Lung images baseline data by deep venous thrombosis diagnosis.

|  | No<br>(N=168) | Yes<br>(N=18) | Overall<br>(N=186) |
| --- | --- | --- | --- |
| **CXR Normal** |  |  |  |
| No | 168 (100%) | 18 (100%) | 186 (100%) |
| Yes | 0 (0%) | 0 (0%) | 0 (0%) |
| **CXR Consolidation** |  |  |  |
| No | 159 (94.6%) | 14 (77.8%) | 173 (93.0%) |
| Yes | 9 (5.4%) | 4 (22.2%) | 13 (7.0%) |
| **CXR Peripheral Lung Interstitial Infiltrates** |  |  |  |
| No | 159 (94.6%) | 15 (83.3%) | 174 (93.5%) |
| Yes | 9 (5.4%) | 3 (16.7%) | 12 (6.5%) |
| **CXR Diffuse Interstitial Lung Disease** |  |  |  |
| No | 148 (88.1%) | 15 (83.3%) | 163 (87.6%) |
| Yes | 20 (11.9%) | 3 (16.7%) | 23 (12.4%) |
| **CXR Pleural Effusion** |  |  |  |
| No | 157 (93.5%) | 16 (88.9%) | 173 (93.0%) |
| Yes | 11 (6.5%) | 2 (11.1%) | 13 (7.0%) |
| **CXR Others** |  |  |  |
| No | 152 (90.5%) | 16 (88.9%) | 168 (90.3%) |
| Yes | 16 (9.5%) | 2 (11.1%) | 18 (9.7%) |
| **CT Normal** |  |  |  |
| No | 167 (99.4%) | 18 (100%) | 185 (99.5%) |
| Yes | 1 (0.6%) | 0 (0%) | 1 (0.5%) |
| **CT Consolidation** |  |  |  |
| No | 84 (50.0%) | 9 (50.0%) | 93 (50.0%) |
| Yes | 84 (50.0%) | 9 (50.0%) | 93 (50.0%) |
| **CT Peripheral Lung Interstitial Infiltrates** |  |  |  |
| No | 161 (95.8%) | 18 (100%) | 179 (96.2%) |
| Yes | 7 (4.2%) | 0 (0%) | 7 (3.8%) |
| **CT Diffuse Interstitial Lung Disease** |  |  |  |
| No | 161 (95.8%) | 18 (100%) | 179 (96.2%) |
| Yes | 7 (4.2%) | 0 (0%) | 7 (3.8%) |
| **CT Ground-Glass** |  |  |  |
| No | 60 (35.7%) | 6 (33.3%) | 66 (35.5%) |
| Yes | 108 (64.3%) | 12 (66.7%) | 120 (64.5%) |
| **CT Pleural Effusion** |  |  |  |
| No | 136 (81.0%) | 14 (77.8%) | 150 (80.6%) |
| Yes | 32 (19.0%) | 4 (22.2%) | 36 (19.4%) |
| **CT Others** |  |  |  |
| No | 127 (75.6%) | 8 (44.4%) | 135 (72.6%) |
| Yes | 41 (24.4%) | 10 (55.6%) | 51 (27.4%) |

CXR = chest X-ray; CT= computed tomography

**Table 4:**
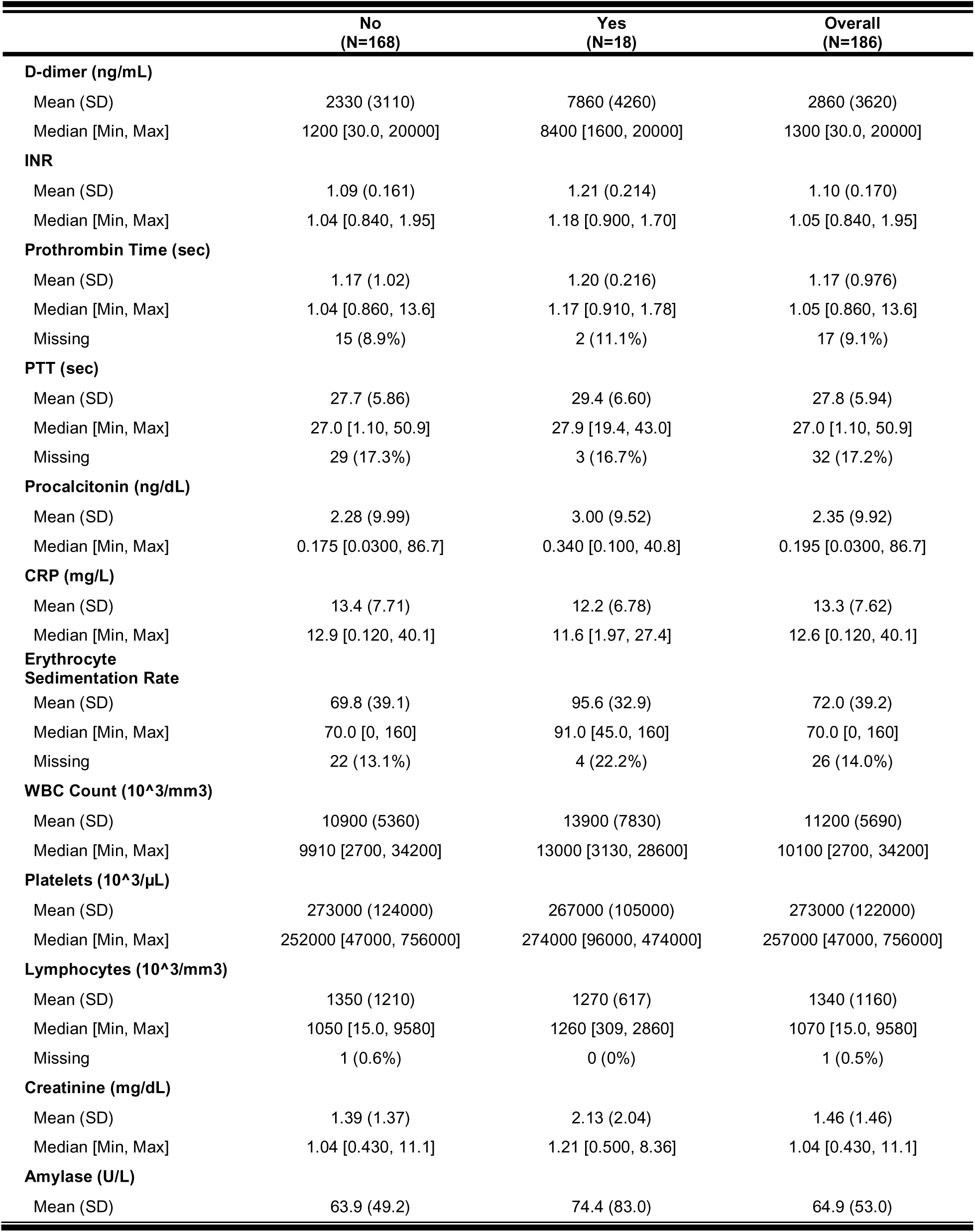
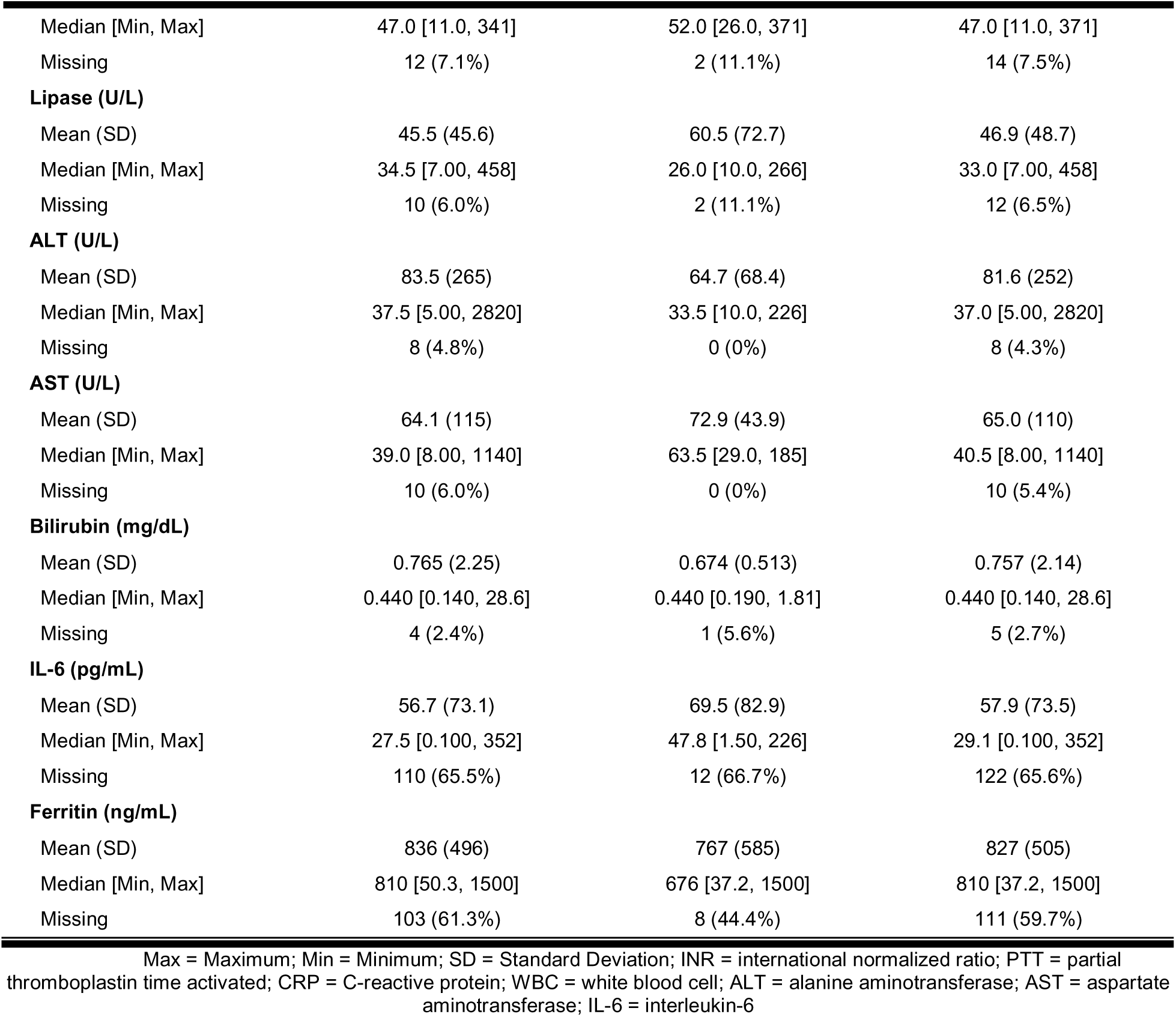
Laboratory baseline data by deep venous thrombosis diagnosis.

The severity was generally higher in the presence of DVT. The CCI and SIC scores showed minimal increases among the participants with DVT compared to the non-DVT group. Additionally, the Wells score was higher in the presence of DVT. (Table 5)

**Table 5:** Physiological and severity scores baseline data by deep venous thrombosis diagnosis.

|  | No<br>(N=168) | Yes<br>(N=18) | Overall<br>(N=186) |
| --- | --- | --- | --- |
| **SOFA** |  |  |  |
| Mean (SD) | 4.26 (3.78) | 6.94 (3.51) | 4.52 (3.83) |
| Median [Min, Max] | 3.00 [0, 13.0] | 8.00 [1.00, 12.0] | 3.00 [0, 13.0] |
| **SAPS 3** |  |  |  |
| Mean (SD) | 48.8 (18.2) | 60.2 (14.6) | 49.9 (18.1) |
| Median [Min, Max] | 44.0 [18.0, 88.0] | 59.5 [40.0, 82.0] | 46.0 [18.0, 88.0] |
| **CHARLSON** |  |  |  |
| Mean (SD) | 2.96 (2.04) | 3.94 (2.29) | 3.06 (2.08) |
| Median [Min, Max] | 3.00 [0, 10.0] | 4.00 [1.00, 8.00] | 3.00 [0, 10.0] |
| **WELLS** |  |  |  |
| Mean (SD) | 0.0774 (0.289) | 3.00 (0.767) | 0.360 (0.938) |
| Median [Min, Max] | 0 [0, 2.00] | 3.00 [2.00, 4.00] | 0 [0, 4.00] |
| **SIC** |  |  |  |
| Mean (SD) | 1.73 (0.887) | 2.56 (0.922) | 1.81 (0.921) |
| Median [Min, Max] | 2.00 [0, 4.00] | 2.00 [1.00, 4.00] | 2.00 [0, 4.00] |

Max = Maximum; Min = Minimum; SAPS3 = Simplified Acute Physiology Score 3; SD = Standard Deviation; SIC = Sepsis-Induced Coagulopathy; SOFA = Sequential Organ Failure Assessment

The Wells Score ranged from zero to four among the participants. The score was sensitive so there were no participants with DVT with a score of zero or one, and also specific as there were no participants without DVT with scores of three or four. The DVT score with highest pair of sensitivity and specificity was two. When considering the score of two or higher to identify with the occurrence of DVT, the sensitivity and specificity confidence intervals were observed to be greater than 80%. (Table 6)

**Table 6:** The diagnostic accuracy of Wells Score performed at admission for detecting DVT among COVID-19 patients.

| Wells Score | D | ND | TP | FN | FP | TN | Sensitivity | Se.inf.ci | Se.sup.ci | Specificity | Sp.inf.ci | Sp.sup.ci |
| --- | --- | --- | --- | --- | --- | --- | --- | --- | --- | --- | --- | --- |
| 0 | 0 | 156 | 18 | 0 | 168 | 1 | 1.000 | 0.824 | 1.000 | 0.006 | 0.001 | 0.033 |
| 1 | 0 | 11 | 18 | 0 | 12 | 156 | 1.000 | 0.824 | 1.000 | 0.929 | 0.879 | 0.959 |
| 2 | 5 | 1 | 18 | 0 | 1 | 167 | 1.000 | 0.824 | 1.000 | 0.994 | 0.967 | 0.999 |
| 3 | 8 | 0 | 13 | 5 | 0 | 168 | 0.722 | 0.491 | 0.875 | 1.000 | 0.978 | 1.000 |
| 4 | 5 | 0 | 5 | 13 | 0 | 168 | 0.278 | 0.125 | 0.509 | 1.000 | 0.978 | 1.000 |

CI = 95% confidence limit; D = With Deep Vein Thrombosis; FN = False Negative; FP = False Positive; ND = Without Deep Vein Thrombosis; Se = Sensitivity; Sp = Specificity; TN = True Negative; TP = True Positive

Hospital mortality and the need for mechanical ventilation were significantly higher among participants with DVT. In contrast, the use of vasopressors was slightly higher in the DVT group, while the length of hospital stay and hemodialysis outcomes were similar among the DVT groups. (Table 7). Additionally, regression models considering the length of stay, vasopressor use, or hemodialysis as outcomes were not significant, either with DVT as a univariate predictor or with DVT adjusted for the SAPS 3 score. (supplemental file)

**Table 7:** Intra-hospital outcomes by deep venous thrombosis diagnosis.

|  | No<br>(N=168) | Yes<br>(N=18) | Overall<br>(N=186) |
| --- | --- | --- | --- |
| **Length of Stay (days)** |  |  |  |
| Mean (SD) | 16.6 (13.8) | 19.8 (16.4) | 16.9 (14.0) |
| Median [Min, Max] | 13.0 [2.00, 87.0] | 15.0 [3.00, 73.0] | 13.0 [2.00, 87.0] |
| **Death** |  |  |  |
| No | 111 (66.1%) | 4 (22.2%) | 115 (61.8%) |
| Yes | 57 (33.9%) | 14 (77.8%) | 71 (38.2%) |
| **Ventilatory Support** |  |  |  |
| No support | 17 (10.1%) | 1 (5.6%) | 18 (9.7%) |
| Inhalatory O2 | 72 (42.9%) | 6 (33.3%) | 78 (41.9%) |
| Facial mask O2 | 47 (28.0%) | 3 (16.7%) | 50 (26.9%) |
| NIV | 4 (2.4%) | 1 (5.6%) | 5 (2.7%) |
| Mechanical Support | 28 (16.7%) | 7 (38.9%) | 35 (18.8%) |
| **Nasal Catheter (L/min)** |  |  |  |
| Mean (SD) | 5.89 (11.7) | 4.75 (3.10) | 5.81 (11.3) |
| Median [Min, Max] | 4.50 [2.00, 89.0] | 4.00 [2.00, 9.00] | 4.50 [2.00, 89.0] |
| Missing | 114 (67.9%) | 14 (77.8%) | 128 (68.8%) |
| **Oxygen Mask (L/min)** |  |  |  |
| Mean (SD) | 9.39 (3.94) | NA (NA) | 9.39 (3.94) |
| Median [Min, Max] | 10.0 [3.00, 15.0] | NA [NA, NA] | 10.0 [3.00, 15.0] |
| Missing | 130 (77.4%) | 18 (100%) | 148 (79.6%) |
| **Norepinephrine** |  |  |  |
| No | 141 (83.9%) | 12 (66.7%) | 153 (82.3%) |
| Yes | 27 (16.1%) | 6 (33.3%) | 33 (17.7%) |
| **Vasopressin** |  |  |  |
| No | 157 (93.5%) | 16 (88.9%) | 173 (93.0%) |
| Yes | 11 (6.5%) | 2 (11.1%) | 13 (7.0%) |
| **Dobutamine** |  |  |  |
| No | 162 (96.4%) | 18 (100%) | 180 (96.8%) |
| Yes | 6 (3.6%) | 0 (0%) | 6 (3.2%) |
| **Nitroglycerin** |  |  |  |
| No | 168 (100%) | 18 (100%) | 186 (100%) |
| Yes | 0 (0%) | 0 (0%) | 0 (0%) |
| **Nitroprusside** |  |  |  |
| No | 160 (95.2%) | 18 (100%) | 178 (95.7%) |
| Yes | 8 (4.8%) | 0 (0%) | 8 (4.3%) |
| **Hemodialysis** |  |  |  |
| No | 148 (88.1%) | 14 (77.8%) | 162 (87.1%) |
| Yes | 20 (11.9%) | 4 (22.2%) | 24 (12.9%) |

Max = Maximum; Min = Minimum; NIV = Noninvasive ventilation; SD = Standard Deviation

DVT was a relevant predictor of both the need for mechanical ventilation (Table 8) and hospital mortality (Table 9). In both analyses, the SAPS 3 severity score was a stronger predictor than DVT. Nevertheless, the presence of DVT explained a relevant amount variation in the survival models and revealed that participants with DVT had twice the hazards compared to participants without DVT, even after adjusting for the SAPS 3 severity score. (Table 8 and Table 9)

**Table 8:** Cox proportional hazards model coefficients and hazard ratios of DVT for the outcome of mechanical ventilation adjusted for SAPS 3 among COVID-19 patients.

| Variables | Effect | S.E. | Lower 0.95 | Upper 0.95 |
| --- | --- | --- | --- | --- |
| **SAPS 3** | 1.494 | 0.227 | 1.050 | 1.938 |
| *Hazard Ratio* | 4.455 |  | 2.857 | 6.945 |
| **Deep Vein Thrombosis - Yes:No** | 0.875 | 0.396 | 0.099 | 1.651 |
| *Hazard Ratio* | 2.399 |  | 1.104 | 5.214 |

Lower = confidence limit; Upper = confidence limit; S.E. = standard error; R2:0.270

**Table 9:** Cox proportional hazards model coefficients and hazard ratios of DVT for mortality adjusted for SAPS 3 among COVID-19 patients.

| Variables | Effect | S.E. | Lower 0.95 | Upper 0.95 |
| --- | --- | --- | --- | --- |
| **SAPS 3** | 1.530 | 0.243 | 1.053 | 2.007 |
| *Hazard Ratio* | 4.619 |  | 2.867 | 7.441 |
| **Deep Vein Thrombosis - Yes: No** | 1.073 | 0.300 | 0.484 | 1.661 |
| *Hazard Ratio* | 2.923 |  | 1.622 | 5.266 |

Lower = confidence limit; Upper = confidence limit; S.E. = standard error; R2:0.262

At a SAPS 3 score as low as twenty, participants with DVT had a survival probability of approximately 55% for remaining free of mechanical ventilation for up to 30 days. Additionally, as the SAPS 3 score increased to 40 and 60, the survival probability decreased to approximately 28 and 5%, respectively, at day thirty for the same participants. (Fig 1)

**Figure 1.**
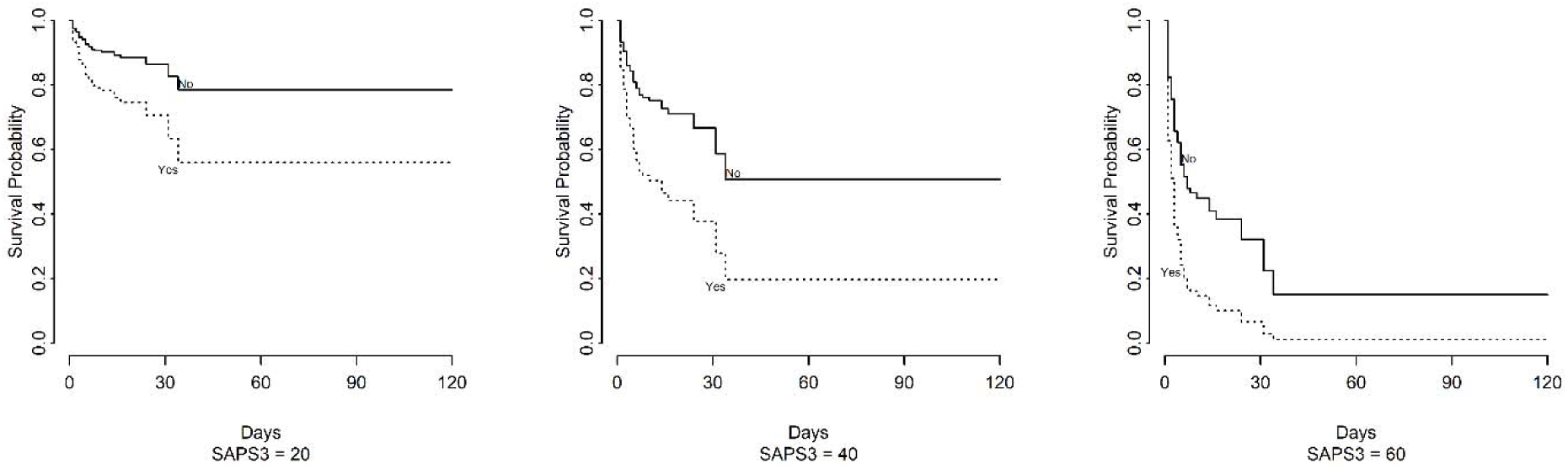
– Cox regression survival curves of patients with and without deep vein thrombosis adjusted for SAPS 3 score taking mechanical ventilation onset as outcome (excluding participants on mechanical ventilation on admission).

At a SAPS 3 score as low as twenty, the survival probability (death outcome) up to 30 days was approximately 80% for the participants with DVT. Additionally, as the SAPS 3 score increased to 40 and 60, the survival probability decreased to approximately 58 and 20%, respectively, at day thirty for the same participants. (Fig 2)

**Figure 2.**
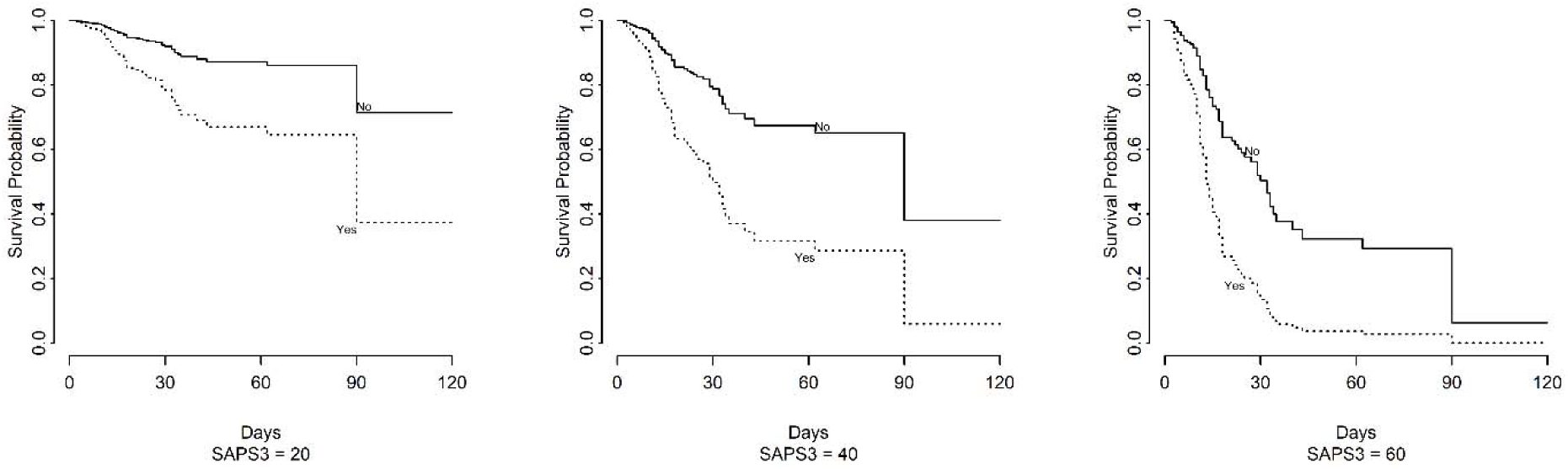
– Cox regression survival curves of patients with and without deep vein thrombosis adjusted for SAPS 3 score taking death as outcome.

In addition, we provide a web calculator available at https://pedrobrasil.shinyapps.io/INDWELL/ to estimate DVT diagnosis probabilities using the Wells score for COVID-19 patients and to estimate the prognosis of critically ill COVID-19 patients with DVT.

## DISCUSSION

The main results to be discussed are as follows: a) the cumulative incidence of DVT and its respective 95% confidence interval was 0.097 [0.062-0.148]. b) The DVT diagnosis by Wells’ score proved to be accurate for COVID-19 patients. c) DVT proved to be a prognostic marker in COVID-19 patients even when adjusted by the SAPS3 severity score.

A few studies have addressed the incidence of venous thromboembolism (VTE) in Coronavirus Disease-2019 (COVID-19) as a phenomenon that still lacks clarity.^39^ Conversely, other researchers assert that there exists a widely accepted consensus regarding the elevated risk of thromboembolic events, such as DVT, associated with SARS-CoV-2 infection.^40^ Nevertheless, several studies estimated the incidence of DVT in critically ill patients with COVID-19 to range from 9.46 to 46%.^2,6–8^ The DVT risk could be as high as 85%. This large variability may be related to the differences in patient sampling, hospital settings, and diagnostic protocols for VTE. Additionally, the risk of DVT appears to be higher in intensive care units despite prophylaxis protocols.^39^

A systematic review of predictive scores for the diagnosis of Pulmonary Embolism in COVID-19 patients published in February 2022 focused on the need to establish new prediction rules, specifically developed and validated for estimating the risk of pulmonary embolism, drawing attention to the limited number of studies on this subject.^41^ Although certain studies have highlighted the incidence of DVT is decreasing with progressive knowledge regarding COVID-19 care, the Wells score was not yet been validated for the population with COVID-19.^42^ However, there is evidence showing it is possible to screen and perform DVT diagnostic investigation in COVID-19 patients using a combination of the Wells score and D-dimer.^43^

An important aspect that strengthens the importance of Wells score validation is the evidence that it is possible to accurately predict DVT diagnosis before the US is performed, and the evidence of DVT as marker of the clinical severity of SARS-CoV-2.

There is association between COVID-19 and VTE. Certain researchers suggest that a cytokine storm is a key factor in fast deterioration of COVID-19, proposing that the knowledge of the time and procedure for blocking the cytokine storm and the time for initiation of anti-inflammatory therapy is critical for reducing the death rate due to COVID-19.^44,45^ The concept of cytokine storm has gradually emerged because of the extremely high mortality associated with multi-organ failure described in the literature. For instance, laboratory experiments conducted on avian Influenza A (H5N1) viruses suggest that virus-induced cytokine dysregulation may contribute to disease severity.^46^

There is a report on acute lung injury and acute respiratory distress syndrome caused partially by host immune responses, suggesting that corticosteroids suppress lung inflammation, which is the case with SARS-CoV infection, as well as influenza, for which systemic inflammation is associated with adverse outcomes.^47^

Hypercoagulability is a hallmark of inflammation. Pro-inflammatory cytokines, such as interleukin (IL)-6, IL-17A, and tumor necrosis factor, are frequently elevated in patients with severe outcomes. Therefore, inflammatory cytokines are critically involved in abnormal clot formation and platelet hyperactivation and play a crucial role in the downregulation of important physiological anticoagulant mechanisms. This suggests that anticoagulant drugs started immediately after confirming SARS-CoV-2 infection to prevent the occurrence of life-threatening complications may be beneficial.^40^

The laboratory tests related to clot disfunction in our study were more frequently abnormal in patients with DVT. The use of anticoagulants in prophylactic doses for venous thromboembolism is indicated for patients hospitalized with COVID-19 according to the Brazilian guidelines from the Ministry of Health. In addition, intermediate doses or therapeutic anticoagulants should not be used in patients with COVID-19 without evidence of thromboembolism.^48^ The COVID-19 Treatment Guidelines issued by the National Institutes of Health (NIH)^49^ recommended the use of a therapeutic dose of heparin for patients with D-dimer levels above the upper normal limit who require low-flow oxygen and who do not have an increased risk of bleeding. However, there is no indication provided by the Brazilian Ministry of Health for the use of D-dimer to guide the use of anticoagulants, nor for the use of anticoagulants after discharge due to COVID-19, even though a segment of the patients may progress to DVT regardless of routine prophylaxis.^48,50,51^ There is an indication for the use of complete anticoagulation therapy in patients with effectively diagnosed atrial fibrillation and venous thromboembolism.^48^ Therefore, the high burden of thrombotic events and their relationship with COVID-19 mortality raises the urgency of this question.^50^

In addition, the American Society of Hematology and the NIH have highlighted the initiation of the use of therapeutic doses of heparin in a non-ICU setting in patients with COVID-19, before transferring them to the ICU. They have also prescribed the use of a therapeutic dose of heparin for patients with D-dimer levels above the upper normal limit who require low-flow oxygen and who do not have an increased risk of bleeding (CIIa). However, it has been advised not to initiate anticoagulation for acutely ill outpatients with COVID-19 and routine anticoagulation for post-discharge patients without a VTE risk. Instead, it is suggested to use scores such as IMPROVEDD to determine whether an individual patient may require post-discharge prophylaxis, considering bleeding risk and feasibility.^52,53^

One of the limitations of this research is the referral involved in the INI/FIOCRUZ hospitalizations. The referral may previously select patients with more severe conditions to be transferred to INI/FICORUZ, as these patients would be more in need to specialized care then other waiting for hospitalization, despite the observed mortality being similar to the mortality reported elsewhere. Nevertheless, if this would be the case, then DVT incidence might be overestimated. At the other hand, as no vaccines were available at the time and mortality rates were high, if transfers were not timely to provide appropriate care, it is possible that severe cases died before they could be observed in the study. If this would be the case, DVT incidence might be underestimated. Nevertheless, the setting was very representative of real-life situation at the time, and this may be an issue when making inferences of DVT incidence to diverse settings.

An additional limitation is that COVID-19 variants of concern changed very rapidly. While the study was going on, the Wild, Zeta and Alpha variants circulated. Later, other variants were preferentially observed. It means that the clinical manifestations and progression to critical illness currently may have other frequencies, and the DVT incidence may require an update in order to inference suit better the current epidemiological setting.

At last, the limitation of the sample size made it hard to detect potential relationship of DVT with some secondary outcomes such as hemodialysis requirement and time using ventilation support.

## CONCLUSIONS

The incidence of DVT in this study was not as high as that reported elsewhere. The evidence suggests that it is possible to accurately diagnose DVT using the Wells score in critically ill patients with COVID-19, regardless of D-dimer test. The Wells score performance was highly accurate and has a potential to replace US in the ICU admission screening, specifically in settings where US is unavailable. DVT increases the severity in critically ill patients with COVID-19 infection, increasing the risk of need for mechanical ventilation, as well as the risk of mortality. Therefore, there is evidence to provide a rationale for guiding anticoagulant prophylaxis and therapy.

## Data Availability

Data may be available from the corresponding author upon reasonable request.

## ACKNOWLEDGEMENTS

The funders had no role in study design, data collection and analysis, decision to publish or preparation of the manuscript. This manuscript was reviewed and edited by Editage (www.editage.com) for English language editing.

All participants or their legal representatives signed an informed consent form before enrollment in RECOVER-SUS-BRASIL. The deep vein thrombosis sub-study protocol was approved by the Ethics Committee INI/FIOCRUZ at October 9^th^, 2020, and can be found at https://plataformabrasil.saude.gov.br/visao/publico/indexPublico.jsf with the number CAEE 32449420.4.1001.5262

The dataset(s) supporting the conclusions of this article will be included at institutional repository at https://www.arca.fiocruz.br/?locale-attribute=en after manuscript publication.

## REFERENCES

1. Ministério da Saúde. Boletim Epidemiológico N° 133 - Boletim COE Coronavírus — Português (Brasil). 2022. Available from: https://www.gov.br/saude/pt-br/centrais-de-conteudo/publicacoes/boletins/epidemiologicos/covid-19/2022/boletim-epidemiologico-no-133-boletim-coe-coronavirus/view [Last accessed: 10/12/2022].

2. Santoliquido A, Porfidia A, Nesci A, et al. Incidence of deep vein thrombosis among non-ICU patients hospitalized for COVID-19 despite pharmacological thromboprophylaxis. J Thromb Haemost 2020;18(9):2358–2363; doi: 10.1111/jth.14992.

3. Li J, Huang DQ, Zou B, et al. Epidemiology of COVID-19: A systematic review and meta-analysis of clinical characteristics, risk factors, and outcomes. J Med Virol 2021;93(3):1449–1458; doi: 10.1002/jmv.26424.

4. Berlin DA, Gulick RM, Martinez FJ. Severe Covid-19. Solomon CG. ed. N Engl J Med 2020;NEJMcp2009575; doi: 10.1056/NEJMcp2009575.

5. Li B, Yang J, Zhao F, et al. Prevalence and impact of cardiovascular metabolic diseases on COVID-19 in China. Clin Res Cardiol 2020; doi: 10.1007/s00392-020-01626-9.

6. Franco-Moreno A, Herrera-Morueco M, Mestre-Gómez B, et al. Incidence of Deep Venous Thrombosis in Patients With COVID-19 and Pulmonary Embolism. J Ultrasound Med 2021;40(7):1411–1416; doi: 10.1002/jum.15524.

7. Baccellieri D, Bertoglio L, Apruzzi L, et al. Incidence of deep venous thrombosis in COVID-19 hospitalized patients during the first peak of the Italian outbreak. Phlebol J Venous Dis 2021;36(5):375–383; doi: 10.1177/0268355520975592.

8. Pieralli F, Pomero F, Giampieri M, et al. Incidence of deep vein thrombosis through an ultrasound surveillance protocol in patients with COVID-19 pneumonia in non-ICU setting: A multicenter prospective study. PLOS ONE 2021;16(5):e0251966; doi: 10.1371/journal.pone.0251966.

9. Cunha MJS, Pinto CAV, Guerra JC de C, et al. Incidence, diagnosis, treatment methods, and outcomes of clinically suspected venous thromboembolic disease in patients with COVID-19 in a quaternary hospital in Brazil. J Vasc Bras 2021;20; doi: 10.1590/1677-5449.200203.

10. Pereira de Godoy JM, Russeff GJ da S, Cunha CH, et al. Increased prevalence of deep vein thrombosis and mortality in patients with Covid-19 at a referral center in Brazil. Phlebology 2022;37(1):21–25; doi: 10.1177/02683555211041931.

11. Pellegrini JAS, Rech TH, Schwarz P, et al. Incidence of venous thromboembolism among patients with severe COVID-19 requiring mechanical ventilation compared to other causes of respiratory failure: a prospective cohort study. J Thromb Thrombolysis 2021;52(2):482– 492; doi: 10.1007/s11239-021-02395-6.

12. Godoy JMP de, Russeff GJDS, Cunha CH, et al. Mortality and Change in the Prevalence of Deep Vein Thrombosis Associated With SARS-CoV-2 P.1 Variant. Cureus 2022;14(7); doi: 10.7759/cureus.26668.

13. Longchamp G, Manzocchi-Besson S, Longchamp A, et al. Proximal deep vein thrombosis and pulmonary embolism in COVID-19 patients: a systematic review and meta-analysis. Thromb J 2021;19(1):15; doi: 10.1186/s12959-021-00266-x.

14. Raskob GE, Spyropoulos AC, Cohen AT, et al. Association Between Asymptomatic Proximal Deep Vein Thrombosis and Mortality in Acutely Ill Medical Patients. J Am Heart Assoc 2021;10(5):e019459; doi: 10.1161/JAHA.120.019459.

15. de Godoy JMP, da Silva MOM, Santos HA, et al. Mortality, deep vein thrombosis, and D-dimer levels in patients with COVID-19. Cor Vasa 2022;64(4):399–402; doi: 10.33678/cor.2022.018.

16. Jasinowodolinski D, Marins Filisbino M, Guedes Baldi B. COVID-19 pneumonia: a risk factor for pulmonary thromboembolism? J Bras Pneumol 2020;46(4):e20200168–e20200168; doi: 10.36416/1806-3756/e20200168.

17. Rizzatti EG, Franco RF. Tratamento do tromboembolismo venoso. Med Ribeirão Preto 2001;34(3/4):269–275; doi: 10.11606/issn.2176-7262.v34i3/4p269-275.

18. COMISSÃO DE CIRCULAÇÃO PULMONAR DA SOCIEDADE BRASILEIRA DE PNEUMOLOGIA E TISIOLOGIA. Recomendações para a prevenção do tromboembolismo venoso. J Pneumol 2000;26:153–158; doi: 10.1590/S0102-35862000000300011.

19. Albricker ACL, Freire CMV, Santos SN dos, et al. Diretriz Conjunta sobre Tromboembolismo Venoso – 2022. Arq Bras Cardiol 2022;118(4):797–857; doi: 10.36660/abc.20220213.

20. Wells PS, Anderson DR, Bormanis J, et al. Value of assessment of pretest probability of deep-vein thrombosis in clinical management. The Lancet 1997;350(9094):1795–1798; doi: 10.1016/S0140-6736(97)08140-3.

21. Bates SM, Jaeschke R, Stevens SM, et al. Diagnosis of DVT: Antithrombotic Therapy and Prevention of Thrombosis, 9th ed: American College of Chest Physicians Evidence-Based Clinical Practice Guidelines. Chest 2012;141(2 Suppl):e351S–e418S; doi: 10.1378/chest.11-2299.

22. Projeto Diretrizes SBACV - Sociedade Brasileira de Angiologia e Cirurgia Vascular. TROMBOSE VENOSA PROFUNDA DIAGNÓSTICO E TRATAMENTO. 2015.

23. Cuker A, Tseng EK, Nieuwlaat R, et al. American Society of Hematology living guidelines on the use of anticoagulation for thromboprophylaxis in patients with COVID-19: May 2021 update on the use of intermediate-intensity anticoagulation in critically ill patients. Blood Adv 2021;5(20):3951–3959; doi: 10.1182/bloodadvances.2021005493.

24. Ren, YAN, Deng, et al. Extremely High Incidence of Lower Extremity Deep Venous Thrombosis in 48 Patients with Severe COVID-19 in Wuhan. /CIRCULATIONAHA 2020.

25. Perazzo H, Cardoso SW, Ribeiro MPD, et al. In-hospital mortality and severe outcomes after hospital discharge due to COVID-19: A prospective multicenter study from Brazil. Lancet Reg Health - Am 2022;11:100244; doi: 10.1016/j.lana.2022.100244.

26. WHO. Emergency Use ICD Codes for COVID-19 Disease Outbreak. 2020. Available from: https://www.who.int/standards/classifications/classification-of-diseases/emergency-use-icd-codes-for-covid-19-disease-outbreak [Last accessed: 10/19/2022].

27. Leonel F. Brasil celebra um ano da vacina contra a Covid-19. 2022. Available from: https://portal.fiocruz.br/noticia/brasil-celebra-um-ano-da-vacina-contra-covid-19 [Last accessed: 10/16/2022].

28. Wells PhilipS, Hirsh J, Anderson DavidR, et al. Accuracy of clinical assessment of deep-vein thrombosis. The Lancet 1995;345(8961):1326–1330; doi: 10.1016/S0140-6736(95)92535-X.

29. Modi S, Deisler R, Gozel K, et al. Wells criteria for DVT is a reliable clinical tool to assess the risk of deep venous thrombosis in trauma patients. World J Emerg Surg WJES 2016;11:24; doi: 10.1186/s13017-016-0078-1.

30. Charlson ME, Carrozzino D, Guidi J, et al. Charlson Comorbidity Index: A Critical Review of Clinimetric Properties. Psychother Psychosom 2022;91(1):8–35; doi: 10.1159/000521288.

31. Raschke RA, Agarwal S, Rangan P, et al. Discriminant Accuracy of the SOFA Score for Determining the Probable Mortality of Patients With COVID-19 Pneumonia Requiring Mechanical Ventilation. JAMA 2021;325(14):1469–1470; doi: 10.1001/jama.2021.1545.

32. Silva Junior JM, Malbouisson LMS, Nuevo HL, et al. Aplicabilidade do escore fisiológico agudo simplificado (SAPS 3) em hospitais brasileiros. Rev Bras Anestesiol 2010;60:20–31; doi: 10.1590/S0034-70942010000100003.

33. Soares M, Silva UVA, Teles JMM, et al. Validation of four prognostic scores in patients with cancer admitted to Brazilian intensive care units: results from a prospective multicenter study. Intensive Care Med 2010;36(7):1188–1195; doi: 10.1007/s00134-010-1807-7.

34. Nassar AP, Malbouisson LS, Moreno R. Evaluation of simplified acute physiology score 3 performance: a systematic review of external validation studies. Crit Care 2014;18(3):R117; doi: 10.1186/cc13911.

35. Pasinato VF, Franzosi OS, Loss SH, et al. SAPS 3 in the modified NUTrition RIsk in the Critically ill score has comparable predictive accuracy to APACHE II as a severity marker. Rev Bras Ter Intensiva 2021;33(3); doi: 10.5935/0103-507X.20210064.

36. Tanaka C, Tagami T, Kudo S, et al. Validation of sepsis-induced coagulopathy score in critically ill patients with septic shock: post hoc analysis of a nationwide multicenter observational study in Japan. Int J Hematol 2021;114(2):164–171; doi: 10.1007/s12185-021-03152-4.

37. Fischer EA, Kinnear B, Sall D, et al. Hospitalist-Operated Compression Ultrasonography: a Point-of-Care Ultrasound Study (HOCUS-POCUS). J Gen Intern Med 2019;34(10):2062–2067; doi: 10.1007/s11606-019-05120-5.

38. Canon Medical Systems. Ultrasound System Transducer Viamo C100. 2020. Available from: https://ch.medical.canon/wp-content/uploads/sites/33/2020/05/Cleaning-guidance-Viamo-c100.pdf [Last accessed: 10/19/2022].

39. Porfidia A, Valeriani E, Pola R, et al. Venous thromboembolism in patients with COVID-19: Systematic review and meta-analysis. Thromb Res 2020;196:67–74; doi: 10.1016/j.thromres.2020.08.020.

40. Tudoran C, Tudoran M, Abu-Awwad A, et al. Spontaneous Hematomas and Deep Vein Thrombosis during the Recovery from a SARS-CoV-2 Infection: Case Report and Literature Review. Medicina (Mex) 2022;58(2):230; doi: 10.3390/medicina58020230.

41. Rindi LV, Al Moghazi S, Donno DR, et al. Predictive scores for the diagnosis of Pulmonary Embolism in COVID-19: A systematic review. Int J Infect Dis 2022;115:93–100; doi: 10.1016/j.ijid.2021.11.038.

42. Cho ES, McClelland PH, Cheng O, et al. Utility of d-dimer for diagnosis of deep vein thrombosis in coronavirus disease-19 infection. J Vasc Surg Venous Lymphat Disord 2021;9(1):47–53; doi: 10.1016/j.jvsv.2020.07.009.

43. Raj K, Chandna S, Doukas SG, et al. Combined Use of Wells Scores and D-dimer Levels for the Diagnosis of Deep Vein Thrombosis and Pulmonary Embolism in COVID-19: A Retrospective Cohort Study. Cureus 2021;13(9); doi: 10.7759/cureus.17687.

44. Zhang S, Li L, Shen A, et al. Rational Use of Tocilizumab in the Treatment of Novel Coronavirus Pneumonia. Clin Drug Investig 2020;40(6):511–518; doi: 10.1007/s40261-020-00917-3.

45. Zhang X, Zhang Y, Qiao W, et al. Baricitinib, a drug with potential effect to prevent SARS-COV-2 from entering target cells and control cytokine storm induced by COVID-19. Int Immunopharmacol 2020;86:106749; doi: 10.1016/j.intimp.2020.106749.

46. de Jong MD, Simmons CP, Thanh TT, et al. Fatal outcome of human influenza A (H5N1) is associated with high viral load and hypercytokinemia. Nat Med 2006;12(10):1203–1207; doi: 10.1038/nm1477.

47. Russell CD, Millar JE, Baillie JK. Clinical evidence does not support corticosteroid treatment for 2019-nCoV lung injury. The Lancet 2020;395(10223):473–475; doi: 10.1016/S0140-6736(20)30317-2.

48. Ministério da Saúde. Diretrizes Brasileiras para Tratamento Hospitalar do Paciente com COVID-19. 2021.

49. NIH. Antithrombotic Therapy in Patients With COVID-19. COVID-19 Treat Guidel 2023.

50. Hanff TC, Mohareb AM, Giri J, et al. Thrombosis in COVID-19. Am J Hematol 2020;95(12):1578–1589; doi: 10.1002/ajh.25982.

51. Brasil M da S Instituto Brasileiro de Geografia e Estatística, Coordenação de Trabalho e Rendimento. Protocolo de Manejo Clínico Da Covid-19 Na Atenção Especializada [Recurso Eletrônico]. Brasilia; 2020.

52. American Society of Hematology. COVID-19 and VTE-Anticoagulation - Hematology.Org. 2022.

53. Russo V, Di Maio M, Attena E, et al. Clinical impact of pre-admission antithrombotic therapy in hospitalized patients with COVID-19: A multicenter observational study. Pharmacol Res 2020;159:104965; doi: 10.1016/j.phrs.2020.104965.

