## supplemental file for "Deep vein thrombosis in critically ill patients with COVID-19: Incidence, Wells score diagnosis validation, and hospital prognosis"

***Table 1****: SIC score diagnostic performance in DVT among Covid-19 patients.*

| SIC Score | D | ND | TP | FN | FP | TN | Sensitivity | Se.inf.cl | Se.sup.cl | Specificity | Sp.inf.cl | Sp.sup.cl |
|---|---|---|---|---|---|---|---|---|---|---|---|---|
| 0 | 0 | 23 | 18 | 0 | 168 | 1 | 1.000 | 0.824 | 1.000 | 0.006 | 0.001 | 0.033 |
| 1 | 1 | 24 | 18 | 0 | 145 | 23 | 1.000 | 0.824 | 1.000 | 0.137 | 0.093 | 0.197 |
| 2 | 10 | 99 | 17 | 1 | 121 | 47 | 0.944 | 0.742 | 0.990 | 0.280 | 0.217 | 0.352 |
| 3 | 3 | 20 | 7 | 11 | 22 | 146 | 0.389 | 0.203 | 0.614 | 0.869 | 0.810 | 0.912 |
| 4 | 4 | 2 | 4 | 14 | 2 | 166 | 0.222 | 0.090 | 0.452 | 0.988 | 0.958 | 0.997 |

cl=95% confidence limit; D=With Deep Vein Thrombosis; FN=False Negative; FP=False Positive; ND=Without Deep Vein Thrombosis; Se=Sensitivity; Sp=Specificity; TN=True Negative; TP=True Positive

***Table 2****: SAPS3 score diagnostic performance in DVT among Covid-19 patients.*

| SAPS3 | D | ND | TP | FN | FP | TN | Sensitivity | Se.inf.cl | Se.sup.cl | Specificity | Sp.inf.cl | Sp.sup.cl |
|---|---|---|---|---|---|---|---|---|---|---|---|---|
| 18 | 0 | 3 | 18 | 0 | 168 | 1 | 1.000 | 0.824 | 1.000 | 0.006 | 0.001 | 0.033 |
| 20 | 0 | 1 | 18 | 0 | 165 | 3 | 1.000 | 0.824 | 1.000 | 0.018 | 0.006 | 0.051 |
| 21 | 0 | 3 | 18 | 0 | 164 | 4 | 1.000 | 0.824 | 1.000 | 0.024 | 0.009 | 0.060 |
| 22 | 0 | 1 | 18 | 0 | 161 | 7 | 1.000 | 0.824 | 1.000 | 0.042 | 0.020 | 0.083 |
| 23 | 0 | 4 | 18 | 0 | 160 | 8 | 1.000 | 0.824 | 1.000 | 0.048 | 0.024 | 0.091 |
| 24 | 0 | 5 | 18 | 0 | 156 | 12 | 1.000 | 0.824 | 1.000 | 0.071 | 0.041 | 0.121 |
| 26 | 0 | 3 | 18 | 0 | 151 | 17 | 1.000 | 0.824 | 1.000 | 0.101 | 0.064 | 0.156 |
| 28 | 0 | 2 | 18 | 0 | 148 | 20 | 1.000 | 0.824 | 1.000 | 0.119 | 0.078 | 0.177 |
| 31 | 0 | 1 | 18 | 0 | 146 | 22 | 1.000 | 0.824 | 1.000 | 0.131 | 0.088 | 0.190 |
| 32 | 0 | 4 | 18 | 0 | 145 | 23 | 1.000 | 0.824 | 1.000 | 0.137 | 0.093 | 0.197 |
| 33 | 0 | 7 | 18 | 0 | 141 | 27 | 1.000 | 0.824 | 1.000 | 0.161 | 0.113 | 0.224 |
| 34 | 0 | 6 | 18 | 0 | 134 | 34 | 1.000 | 0.824 | 1.000 | 0.202 | 0.149 | 0.269 |
| 35 | 0 | 2 | 18 | 0 | 128 | 40 | 1.000 | 0.824 | 1.000 | 0.238 | 0.180 | 0.308 |
| 36 | 0 | 9 | 18 | 0 | 126 | 42 | 1.000 | 0.824 | 1.000 | 0.250 | 0.191 | 0.321 |
| 37 | 0 | 1 | 18 | 0 | 117 | 51 | 1.000 | 0.824 | 1.000 | 0.304 | 0.239 | 0.377 |
| 38 | 0 | 10 | 18 | 0 | 116 | 52 | 1.000 | 0.824 | 1.000 | 0.310 | 0.245 | 0.383 |
| 39 | 0 | 1 | 18 | 0 | 106 | 62 | 1.000 | 0.824 | 1.000 | 0.369 | 0.300 | 0.444 |
| 40 | 1 | 1 | 18 | 0 | 105 | 63 | 1.000 | 0.824 | 1.000 | 0.375 | 0.305 | 0.450 |
| 41 | 0 | 5 | 17 | 1 | 104 | 64 | 0.944 | 0.742 | 0.990 | 0.381 | 0.311 | 0.456 |
| 42 | 1 | 3 | 17 | 1 | 99 | 69 | 0.944 | 0.742 | 0.990 | 0.411 | 0.339 | 0.486 |
| 43 | 1 | 5 | 16 | 2 | 96 | 72 | 0.889 | 0.672 | 0.969 | 0.429 | 0.356 | 0.504 |
| 44 | 1 | 8 | 15 | 3 | 91 | 77 | 0.833 | 0.608 | 0.942 | 0.458 | 0.385 | 0.534 |
| 45 | 1 | 2 | 14 | 4 | 83 | 85 | 0.778 | 0.548 | 0.910 | 0.506 | 0.431 | 0.581 |
| 46 | 1 | 3 | 13 | 5 | 81 | 87 | 0.722 | 0.491 | 0.875 | 0.518 | 0.443 | 0.592 |
| 47 | 0 | 2 | 12 | 6 | 78 | 90 | 0.667 | 0.437 | 0.837 | 0.536 | 0.460 | 0.609 |
| 48 | 0 | 7 | 12 | 6 | 76 | 92 | 0.667 | 0.437 | 0.837 | 0.548 | 0.472 | 0.621 |
| 49 | 0 | 2 | 12 | 6 | 69 | 99 | 0.667 | 0.437 | 0.837 | 0.589 | 0.514 | 0.661 |
| 50 | 0 | 1 | 12 | 6 | 67 | 101 | 0.667 | 0.437 | 0.837 | 0.601 | 0.526 | 0.672 |
| 52 | 0 | 1 | 12 | 6 | 66 | 102 | 0.667 | 0.437 | 0.837 | 0.607 | 0.532 | 0.678 |
| 53 | 0 | 2 | 12 | 6 | 65 | 103 | 0.667 | 0.437 | 0.837 | 0.613 | 0.538 | 0.683 |
| 54 | 0 | 3 | 12 | 6 | 63 | 105 | 0.667 | 0.437 | 0.837 | 0.625 | 0.550 | 0.695 |
| 55 | 0 | 1 | 12 | 6 | 60 | 108 | 0.667 | 0.437 | 0.837 | 0.643 | 0.568 | 0.711 |

| SAPS3 | D | ND | TP | FN | FP | TN | Sensitivity | Se.inf.cl | Se.sup.cl | Specificity | Sp.inf.cl | Sp.sup.cl |
|---|---|---|---|---|---|---|---|---|---|---|---|---|
| 56 | 1 | 7 | 12 | 6 | 59 | 109 | 0.667 | 0.437 | 0.837 | 0.649 | 0.574 | 0.717 |
| 57 | 2 | 0 | 11 | 7 | 52 | 116 | 0.611 | 0.386 | 0.797 | 0.690 | 0.617 | 0.755 |
| 58 | 0 | 3 | 9 | 9 | 52 | 116 | 0.500 | 0.290 | 0.710 | 0.690 | 0.617 | 0.755 |
| 59 | 0 | 1 | 9 | 9 | 49 | 119 | 0.500 | 0.290 | 0.710 | 0.708 | 0.636 | 0.772 |
| 62 | 2 | 2 | 9 | 9 | 48 | 120 | 0.500 | 0.290 | 0.710 | 0.714 | 0.642 | 0.777 |
| 63 | 0 | 3 | 7 | 11 | 46 | 122 | 0.389 | 0.203 | 0.614 | 0.726 | 0.654 | 0.788 |
| 64 | 0 | 2 | 7 | 11 | 43 | 125 | 0.389 | 0.203 | 0.614 | 0.744 | 0.673 | 0.804 |
| 65 | 0 | 4 | 7 | 11 | 41 | 127 | 0.389 | 0.203 | 0.614 | 0.756 | 0.686 | 0.815 |
| 66 | 1 | 2 | 7 | 11 | 37 | 131 | 0.389 | 0.203 | 0.614 | 0.780 | 0.711 | 0.836 |
| 67 | 0 | 2 | 6 | 12 | 35 | 133 | 0.333 | 0.163 | 0.563 | 0.792 | 0.724 | 0.846 |
| 68 | 1 | 3 | 6 | 12 | 33 | 135 | 0.333 | 0.163 | 0.563 | 0.804 | 0.737 | 0.857 |
| 69 | 0 | 1 | 5 | 13 | 30 | 138 | 0.278 | 0.125 | 0.509 | 0.821 | 0.757 | 0.872 |
| 71 | 0 | 1 | 5 | 13 | 29 | 139 | 0.278 | 0.125 | 0.509 | 0.827 | 0.763 | 0.877 |
| 72 | 0 | 1 | 5 | 13 | 28 | 140 | 0.278 | 0.125 | 0.509 | 0.833 | 0.770 | 0.882 |
| 73 | 0 | 4 | 5 | 13 | 27 | 141 | 0.278 | 0.125 | 0.509 | 0.839 | 0.776 | 0.887 |
| 74 | 0 | 3 | 5 | 13 | 23 | 145 | 0.278 | 0.125 | 0.509 | 0.863 | 0.803 | 0.907 |
| 75 | 0 | 1 | 5 | 13 | 20 | 148 | 0.278 | 0.125 | 0.509 | 0.881 | 0.823 | 0.922 |
| 76 | 0 | 4 | 5 | 13 | 19 | 149 | 0.278 | 0.125 | 0.509 | 0.887 | 0.830 | 0.926 |
| 78 | 3 | 3 | 5 | 13 | 15 | 153 | 0.278 | 0.125 | 0.509 | 0.911 | 0.858 | 0.945 |
| 79 | 1 | 0 | 2 | 16 | 12 | 156 | 0.111 | 0.031 | 0.328 | 0.929 | 0.879 | 0.959 |
| 80 | 0 | 2 | 1 | 17 | 12 | 156 | 0.056 | 0.010 | 0.258 | 0.929 | 0.879 | 0.959 |
| 81 | 0 | 1 | 1 | 17 | 10 | 158 | 0.056 | 0.010 | 0.258 | 0.940 | 0.894 | 0.967 |
| 82 | 1 | 6 | 1 | 17 | 9 | 159 | 0.056 | 0.010 | 0.258 | 0.946 | 0.901 | 0.972 |
| 83 | 0 | 1 | 0 | 18 | 3 | 165 | 0.000 | 0.000 | 0.176 | 0.982 | 0.949 | 0.994 |
| 88 | 0 | 2 | 0 | 18 | 2 | 166 | 0.000 | 0.000 | 0.176 | 0.988 | 0.958 | 0.997 |

cl=95% confidence limit; D=With Deep Vein Thrombosis; FN=False Negative; FP=False Positive; ND=Without Deep Vein Thrombosis; Se=Sensitivity; Sp=Specificity; TN=True Negative; TP=True Positive

***Table 3**: Charlson score diagnostic performance in DVT among Covid-19 patients.*

| Charlson Score | D | ND | TP | FN | FP | TN | Sensitivity | Se.inf.cl | Se.sup.cl | Specificity | Sp.inf.cl | Sp.sup.cl |
|---|---|---|---|---|---|---|---|---|---|---|---|---|
| 0 | 0 | 24 | 18 | 0 | 168 | 1 | 1.000 | 0.824 | 1.000 | 0.006 | 0.001 | 0.033 |
| 1 | 3 | 17 | 18 | 0 | 144 | 24 | 1.000 | 0.824 | 1.000 | 0.143 | 0.098 | 0.204 |
| 2 | 2 | 38 | 15 | 3 | 127 | 41 | 0.833 | 0.608 | 0.942 | 0.244 | 0.185 | 0.314 |
| 3 | 3 | 24 | 13 | 5 | 89 | 79 | 0.722 | 0.491 | 0.875 | 0.470 | 0.396 | 0.546 |
| 4 | 4 | 21 | 10 | 8 | 65 | 103 | 0.556 | 0.337 | 0.754 | 0.613 | 0.538 | 0.683 |
| 5 | 3 | 23 | 6 | 12 | 44 | 124 | 0.333 | 0.163 | 0.563 | 0.738 | 0.667 | 0.799 |
| 6 | 0 | 18 | 3 | 15 | 21 | 147 | 0.167 | 0.058 | 0.392 | 0.875 | 0.816 | 0.917 |
| 8 | 3 | 2 | 3 | 15 | 3 | 165 | 0.167 | 0.058 | 0.392 | 0.982 | 0.949 | 0.994 |
| 10 | 0 | 1 | 0 | 18 | 1 | 167 | 0.000 | 0.000 | 0.176 | 0.994 | 0.967 | 0.999 |

cl=95% confidence limit; D=With Deep Vein Thrombosis; FN=False Negative; FP=False Positive; ND=Without Deep Vein Thrombosis; Se=Sensitivity; Sp=Specificity; TN=True Negative; TP=True Positive

***Table 4**: SOFA score diagnostic performance in DVT among Covid-19 patients.*

| SOFA Score | D | ND | TP | FN | FP | TN | Sensitivity | Se.inf.cl | Se.sup.cl | Specificity | Sp.inf.cl | Sp.sup.cl |
|---|---|---|---|---|---|---|---|---|---|---|---|---|
| 0 | 0 | 23 | 18 | 0 | 168 | 1 | 1.000 | 0.824 | 1.000 | 0.006 | 0.001 | 0.033 |
| 1 | 1 | 25 | 18 | 0 | 145 | 23 | 1.000 | 0.824 | 1.000 | 0.137 | 0.093 | 0.197 |
| 2 | 3 | 32 | 17 | 1 | 120 | 48 | 0.944 | 0.742 | 0.990 | 0.286 | 0.223 | 0.358 |
| 3 | 0 | 18 | 14 | 4 | 88 | 80 | 0.778 | 0.548 | 0.910 | 0.476 | 0.402 | 0.551 |
| 4 | 1 | 9 | 14 | 4 | 70 | 98 | 0.778 | 0.548 | 0.910 | 0.583 | 0.508 | 0.655 |
| 5 | 1 | 5 | 13 | 5 | 61 | 107 | 0.722 | 0.491 | 0.875 | 0.637 | 0.562 | 0.706 |
| 6 | 1 | 6 | 12 | 6 | 56 | 112 | 0.667 | 0.437 | 0.837 | 0.667 | 0.592 | 0.734 |
| 7 | 1 | 4 | 11 | 7 | 50 | 118 | 0.611 | 0.386 | 0.797 | 0.702 | 0.629 | 0.766 |
| 8 | 3 | 9 | 10 | 8 | 46 | 122 | 0.556 | 0.337 | 0.754 | 0.726 | 0.654 | 0.788 |
| 9 | 1 | 17 | 7 | 11 | 37 | 131 | 0.389 | 0.203 | 0.614 | 0.780 | 0.711 | 0.836 |
| 10 | 4 | 8 | 6 | 12 | 20 | 148 | 0.333 | 0.163 | 0.563 | 0.881 | 0.823 | 0.922 |
| 11 | 1 | 3 | 2 | 16 | 12 | 156 | 0.111 | 0.031 | 0.328 | 0.929 | 0.879 | 0.959 |
| 12 | 1 | 8 | 1 | 17 | 9 | 159 | 0.056 | 0.010 | 0.258 | 0.946 | 0.901 | 0.972 |
| 13 | 0 | 1 | 0 | 18 | 1 | 167 | 0.000 | 0.000 | 0.176 | 0.994 | 0.967 | 0.999 |

cl=95% confidence limit; D=With Deep Vein Thrombosis; FN=False Negative; FP=False Positive; ND=Without Deep Vein Thrombosis; Se=Sensitivity; Sp=Specificity; TN=True Negative; TP=True Positive

***Table 5**: Least square linear model DVT and SAPS 3 coefficients predicting log length of stay among Covid-19 patients.*

| Variables | Effect | S.E. | Lower 0.95 | Upper 0.95 |
|---|---|---|---|---|
| SAPS 3 | 0.312 | 0.081 | 0.153 | 0.471 |
| TVP - Yes:No | 0.059 | 0.170 | -0.276 | 0.395 |

Lower = confidence limit; Upper = confidence limit; S.E. = standard error;
R2 ajusted: 0.071

***Table 6**: Least square linear model DVT and SAPS 3 coefficients predicting log length of mechanical ventilation among Covid-19 patients.*

| Variables | Effect | S.E. | Lower 0.95 | Upper 0.95 |
|---|---|---|---|---|
| SAPS 3 | 0.346 | 0.133 | 0.083 | 0.610 |
| TVP - Yes:No | -0.445 | 0.231 | -0.904 | 0.015 |

Lower = confidence limit; Upper = confidence limit; S.E. = standard error;
R2 ajusted: 0.075
